# Intestinal inflammation induced by heat-labile toxin-producing Enterotoxigenic *E. Coli* infection and impact on immune responses in an experimental human challenge model

**DOI:** 10.1101/2025.04.03.25325163

**Authors:** Xueyan Zhang, Jessica Brubaker, Kawsar R. Talaat, Chad K. Porter, Brittany L Feijoo, Brittany M Adjoodani, Barbara DeNearing, Michael G Prouty, A Louis Bourgeois, David A Sack, Susanne Eder-Lingelbach, Christian Taucher, Subhra Chakraborty

## Abstract

Enterotoxigenic *Escherichia coli* (ETEC) causes significant morbidity, mortality, and growthth faltering among children, particularly in low- and middle-income countries. While gut inflammation contributes to growth faltering, the role of ETEC in inflammation remains poorly understood. We previously demonstrated that ETEC-producing heat-labile toxin (LT) and heat-stable toxins (ST) induced significant inflammation in humans, but LT-only strains are understudied. In this study, we evaluated the intestinal inflammation induced by the LT-only ETEC strain LSN03-016011/A in a human challenge model. Stool samples were analyzed for pre- and post-challenge myeloperoxidase (MPO) and pro and anti-inflammatory cytokines, ETEC shedding, and ETEC-specific antibody responses. MPO, IL-1β, and IL-8 levels significantly increased post-ETEC challenge, but there was no significant difference between symptomatic and asymptomatic participants. Participants protected from severe diarrhea had higher levels of pre-challenge IL-10, IL-13, and IFN-γ compared to those not protected. The MPO and specific cytokine levels were significantly correlated with the seroconversion status to LT and the colonization factor antigen CS17. This study provides evidence that LT-ETEC strain can induce significant intestinal inflammation even in the absence of symptoms, highlighting the need for a vaccine and a better understanding of the impact of ETEC-attributable inflammation on child health in endemic areas.

**Author summary:** Enterotoxigenic *Escherichia coli* (ETEC) is one of the leading causes of enteric infections, resulting in diarrhea, malnutrition, and other long-term health effects. However, how ETEC - particularly strains that produce only the heat-labile toxin (LT) - can contribute to gut inflammation is not fully understood. In this study, we examined the impact of an LT-ETEC infection on gut inflammation and its relations to ETEC-specific immune responses using samples from participants in a controlled human infection study. We found that LT-ETEC induces a significant level of gut inflammation marker myeloperoxidase (MPO) and pro-inflammatory cytokines IL-8 and IL-1β, when the patients had moderate to severe diarrhea and even when diarrheal symptoms were mild or absent. Gut inflammation level correlated with immune responses to ETEC. These findings suggest that LT-ETEC infection causes significant gut inflammation, which plays a role in immune responses. Our results highlight the need for preventive strategies to reduce the burden of ETEC-related illness, particularly in regions where these infections are common, to prevent broader adverse consequences for gut health and child development.

## Introduction

Enterotoxigenic *Escherichia coli* (ETEC) is an important bacterial pathogen causing diarrhea in children in low and middle-income countries. Initial ETEC infections can be early in life^1^, and were estimated to cause 222 million cases of diarrhea in 2016, including 75 million in children under five years old^2^. The annual mortality from the illness due to ETEC was estimated at 51,186, 36% of which was among children under 5, and 88% in Sub-Saharan Africa and South Asia^2^. Another estimate in 2010 projected 8.5 million ETEC Disability-Adjusted Life Years and 1 million Years Lived with Disability^3,4^. Adults between the ages of 20 and 60 years are also at high risk of ETEC infection^5,6^, but limited data are available. In addition, ETEC remains the leading cause of infectious diarrhea among international travelers, and military service members deployed to the ETEC endemic areas^7,8^. Besides immediate clinical outcomes, ETEC infections are associated with long-term adverse consequences, including growth faltering^9,10^, potentially driven by inflammation due to enteric pathogens^11–13^.

Inflammation caused by ETEC infection has previously been described^12^. In murine and porcine models, ETEC increased MPO, lipocalin-2 (LCN-2), IL-1β, IL-6, IL-8, IL-17, IL-18 and TNF-α, and decreased IL-4 and TGF-β^14–21^. In humans, ETEC infection was associated with increased MPO in the Malnutrition and Enteric Disease Study (MAL-ED)^8,22^. Additionally, ETEC was significantly associated with increased fecal calprotectin in Bangladeshi children^23^. However, critical knowledge gaps still exist. Studies in cell culture and/or animal models may not fully reflect the impact on humans. Additionally, most human studies are observational and conducted in endemic areas, making causality difficult to assess, particularly in the presence of co-pathogens. Also, questions remain about whether infections with all ETEC pathotypes contribute equally to the inflammatory pathways, which could lead to deficits in human physical and cognitive development. In the Global Enteric Multicenter Study (GEMS) study, ETEC strains that produced only heat-labile toxin (LT) were found equally prevalent among the cases that needed urgent care and among the asymptomatic children and, therefore, were not significantly attributed to the acute diarrhea cases as opposed to ETEC strains that expressed LT and heat stable toxin (ST) as well as those that produced only ST^24^.

The controlled human infection model (CHIM) has the advantage of evaluating the impact of ETEC infection on host inflammation, using well-characterized strains. We previously demonstrated that ETEC strain H10407, which co-expresses LT and ST, induced significant systemic and intestinal inflammation among both symptomatic and asymptomatic volunteers in a CHIM^25^. Similarly, two strains of ST-only ETEC have been shown to increase systemic inflammation and mucosal damage, even among asymptomatic patients^26^.

However, the extent to which ETEC strains producing only LT (LT-ETEC) induce inflammation in humans has not been widely studied. LT facilitates ETEC colonization and is a key component in ETEC vaccine candidates due to its ability to induce an anti-LT antibody response^27^. Recent data in animal models also indicate that LT drives several enteropathic changes in the small intestinal epithelia that can contribute to long-term negative health consequences and modulate other cellular changes and intestinal receptor expression that impact susceptibility to ETEC and other enteric pathogens^28–32^. In addition, LT is essential to activating both NF-κB and MAPK signaling pathways and inducing proinflammatory cytokines and MPO, as shown in animal models^33, 24^. On the other hand, an *in vitro* study demonstrated that LT impaired macrophage-mediated phagocytosis of ETEC, thereby hindering the initiation of inflammatory responses to the pathogen^34^.

In this study, we evaluated the intestinal inflammation induced by an LT-ETEC strain in a human challenge model. We measured the kinetics of MPO and a panel of pro- and anti-inflammatory cytokines in the stool of the volunteers challenged with the LT-ETEC strain. We also evaluated any impact of ETEC-induced intestinal inflammation on ETEC colonization (shedding in stool), ETEC disease severity, and immune responses to ETEC vaccine-specific antigens, LTB and CS17.

## Materials and methods

### Experimental human challenge model of LT-ETEC

Samples were obtained from a single-center experimental challenge study among 15 American adults challenged with 5×10^9^ CFU of ETEC strain LSN03-016011/A, producing LT and the colonization factor CS17^35^. The study was approved by the Bloomberg School of Public Health, Johns Hopkins University IRB (#00008616), and conducted in the in-patient unit at the Center for Immunization Research, Johns Hopkins University. Volunteers were ineligible for this study if they received another investigational product within 30 days; were exposed to ETEC or had a history of diarrhea when traveling in developing countries within the past three years; had medical concerns, abnormal stool patterns, chronic diseases, HIV-1/hepatitis B/hepatitis C infection, allergies to antibiotics, a recent history of alcohol/drug abuse, or were pregnant.

The challenge occurred on Day 1, with all Day 1 measures conducted post-challenge. Day 0 values (1 day before the challenge) of MPO, cytokines, and antibody titers were identified as baseline values. Moderate to severe diarrhea (MSD) was defined as at least 4 loose stools or >400 g in 24 hours. Mild diarrhea was defined as 1-3 loose stools or loose stools with ≤ 400 g in 24 hours. All participants received antibiotic treatment 120 hours after the challenge or when they met the criteria for early treatment. Stool samples were collected before and every day following the ETEC challenge. If a stool sample was not available, a rectal swab was obtained at least once per day. Up to 3 stools or rectal swabs per day post-challenge were assessed by culture daily until two consecutive samples were negative for ETEC, using methods previously described^36–38^. Quantitative stool culture was performed 2 and 4 days post-challenge.^36–38^ Blood samples were collected before the challenge and 7 and 28 days post-challenge. All samples were stored at - 80°C after collection. The diarrhea severity score was calculated as previously described^39^.

### Evaluation of inflammation biomarkers

MPO levels in stool were measured by ELISA (Immundiagnostik, cat# K 6603) as per the manufacturer’s instructions. Stool samples were diluted 1:500 with the wash buffer, and 100μl of the diluted stool samples were used for the assay. Samples, detection antibody, conjugate, substrate, and stop solution were added in order, each followed by incubation and washing. Results were calculated using a standard curve and adjusted by the weight of the stool.

The levels of 10 cytokines, including IL-1β, IL-2, IL-4, IL-6, IL-8, IL-10, IL-13, IL-17A, TNF-α, and IFN-γ were tested in stool samples with Meso-Scale Discovery multispot array (MSD, Gaithersburg, MD). The levels of cytokines were read with an MS2400 imager (MSD). The lowest limit of quantification (LLOQ) was defined as the lowest calibrator value with the coefficient of variation ≤ 25% and recovery of the calibrator ≤ 25% of the expected value^38^. Cytokine levels were considered undetectable if they were below the LLOQ and were assigned as the value of LLOQ for analysis.

### Evaluation of immune responses

Immune responses including IgA and IgG in serum and IgA in lymphocyte supernatant (ALS) and fecal samples against CS17 and CTB were measured by ELISA^36,40,41^. CTB (Cholera Toxin Subunit B) is structurally and immunogenically similar to LTB and is used as a proxy. Samples were serially diluted in 3-fold increments and tested in duplicate. Samples from the same participant were tested on a single plate, with controls included on each plate for consistency. Goat anti-human IgG or IgA conjugated to horseradish peroxidase (HRP) (KPL, Gaithersburg, MD) was used as the secondary antibody. The titer was calculated as the reciprocal dilution that produced an optical density of 0.2 for serum samples and 0.4 for ALS and fecal samples above the background at 450 nm. The final titer was adjusted by multiplying the ratio of the geometric mean of all samples on each plate divided by the geometric mean of all positive controls across plates.

### Statistical analysis

Data were analyzed using R (version 4.0.2)^42^, and figures were created using GraphPad Prism (GraphPad, CA). Peak values were identified as the highest titers of variables on any day following the challenge. Peak fold change was calculated as the ratio of peak value to baseline value. The geometric mean (GM) values across participants on any day were calculated for MPO and cytokines. Peak GM values were determined by calculating the GM across participants on any day and identifying the highest value. The GM of peak values was determined by taking all peak values and calculating the GM across participants. At least a 2-fold change in the GM of cytokines was considered to have clinical significance. A responder was defined as a >= 2-fold increase from baseline in the IgA and IgG titer in serum and a >= 4-fold increase in ALS and stool^36–38^. Nonparametric analyses were employed due to the small sample size. Comparisons were conducted with Wilcoxon rank sum tests for MPO and cytokines to evaluate differences between participants with MSD and those without, as well as between those who seroconverted and those who did not. Wilcoxon matched pairs test was applied to assess changes in MPO and cytokine levels on each day following the challenge relative to baseline or peak values. The levels of LT-ETEC-induced MPO were compared with the previously published (LT+ST)-ETEC (H10407 strain) induced MPO^25^. Spearman correlation was used to quantify the correlation between MPO, cytokines, ETEC shedding, and disease severity score. Spearman’s rank correlation coefficient (ρ) > 0.7 or < -0.7 was considered to indicate a strong correlation, and a p value < 0.05 was considered significant in the analysis^43^.

## Results

### Clinical outcomes

Among the 15 naïve participants who were challenged with ETEC, 11 participants developed MSD, while 4 experienced mild diarrhea (3) or no diarrhea (1), resulting in a MSD rate of 73.3%. For analysis in this study, the participants with mild and no diarrhea are grouped as non-MSD (no moderate to severe disease).

No significant differences were found between those with and without MSD in terms of age, gender, race, or ethnicity. The median age at enrollment was 34.0 years for those with MSD and 33.5 years for those without (p = 1). 5 participants (45.5%) with MSD were male, compared to 3 participants (75%) without MSD (p = 0.668). Regarding race and ethnicity, among those with MSD, 9 participants were Black or African American (one of whom was Hispanic or Latino) and 2 were White. Among those without MSD, 2 participants were Black or African American, 1 identified as another racial group, and 1 was Hispanic or Latino.

Participants with MSD had significantly higher disease severity scores (p=0.007), a greater maximum number (p=0.005) and volume of loose stools in a 24-hour period (p=0.006), and a higher total number (p=0.011) and volume of loose stools (p=0.010), compared to those without MSD (Table 1). A total of 8 (53.3%) participants received early antibiotic treatment.

**Table 1.** Clinical outcomes of participants challenged with ETEC.

|  | Overall<br>(N=15) | MSD<br>(N=11) | non-MSD<br>(N=4) | p |
| --- | --- | --- | --- | --- |
| Disease severity score | 2.0 [1.0;3.0] | 3.0 [2.0; 3.0] | 1.0 [0.5; 1.0] | 0.007 |
| Duration of diarrhea (Hours) | 44.6 [24.1;52.4] | 44.6 [31.2;52.4] | 25.8 [3.4;52.1] | 0.489 |
| Max number in 24h (any grade) | 5.0 [4.0;7.0] | 6.0 [5.0; 7.5] | 3.0 [3.0; 3.5] | 0.006 |
| Max number in 24h (loose/liquid stool) | 5.0 [3.5;7.0] | 6.0 [5.0; 7.5] | 2.0 [1.0; 2.5] | 0.005 |
| Max volume in 24h (loose/liquid stool) | 521.0 [268.0;887.0] | 855.0 [463.0;952.0] | 105.5 [16.5;268.0] | 0.006 |
| Time to onset of diarrhea from<br>receiving the challenge (Hours) | 20.0 [10.8;30.6] | 21.9 [11.1;29.1] | 16.7 [12.6;33.9] | 1.000 |
| Time to onset of MSD from receiving<br>the challenge (Hours) | 26.4 [14.7;41.0] | 26.4 [14.7;41.0] |  |  |
| Total number of loose stools | 9.0 [7.0;11.5] | 10.0 [9.0;13.0] | 3.0 [1.0; 6.0] | 0.011 |
| Total volume of loose stools (mL) | 855.0 [477.0;1401.0] | 1226.0 [793.0;1595.0] | 172.0 [16.5;480.0] | 0.010 |
| Time to antibiotic treatment (Hours) | 81.5 [42.6;120.0] | 59.8 [37.8;100.8] | 120.0 [120.0;120.0] | 0.033 |

Data are shown as median[IQR] and compared with wilcoxon test.

### Kinetics of intestinal inflammation biomarker MPO after challenge

Fecal MPO levels increased significantly on days 2-6 and day 10 following ETEC challenge, peaking at day 3 with a geometric mean (GM) of 3602.00 ng/ml (95% CI: 1952.54, 6489.71), a 10.63-fold rise from baseline (338.95 ng/ml; 95% CI: 266.78, 437.36; p<0.001) (Fig 1A). MPO decreased after receiving antibiotic treatment, with significant reductions on day 7 and day 8 compared to day 6 (p=0.002, p=0.016).

**Fig 1.**
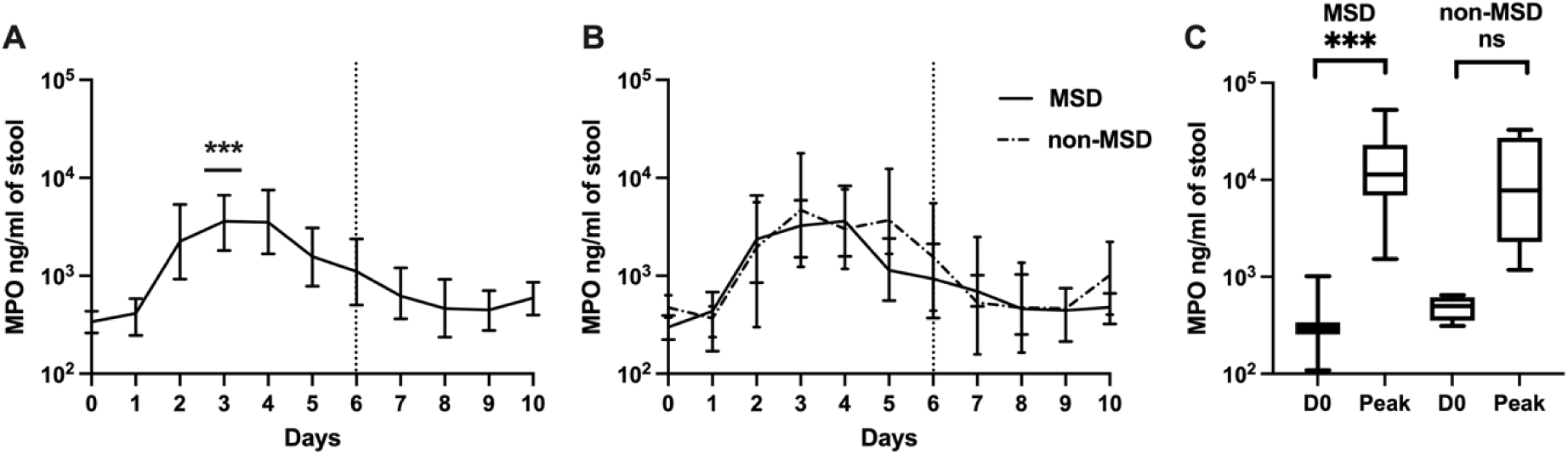
MPO levels pre- and post-ETEC challenge. (log 10 scale).

Participants with and without MSD showed a similar MPO trend, with an increase after the challenge and a decrease after antibiotic treatment (Fig 1B). Among participants with MSD, MPO peaked at day 4 with a GM of 3629.38 ng/ml (95% CI: 1522.31, 8495.06), a 12.10-fold rise from baseline (p=0.002). In non-MSD participants, MPO peaked at day 3 with a GM of 4688.85 ng/ml (95% CI: 1232.88, 17832.49), a 9.89-fold increase.

The GM peak MPO on any day across all participants was 9397.35 (95%CI: 5416.25, 16549.90) ng/ml, 27.73-fold higher than the baseline (p<0.001). Similarly, the GM of peak MPO was 35.21 (p<0.001) fold higher among those with MSD and 14.37 (p=0.125) fold higher among those without MSD compared to the baseline (Fig 1C). Baseline and peak MPO levels did not differ significantly between those with and without MSD (baseline: non-MSD/MSD=1.58; peak: MSD/non-MSD=1.55; p>0.05).

A. Magnitude and kinetics of MPO levels among all participants. B. Magnitude and kinetics of MPO stratified by clinical outcome following ETEC challenge. C. Comparison of baseline MPO and peak post-challenge MPO levels by clinical outcome. D0: day before challenge; Peak: peak MPO levels on any day. MSD: moderate to severe diarrhea; non-MSD: without moderate to severe diarrhea. p<0.001: ***; not significant: NS. The dashed vertical line on Day 6 (120h post-challenge) in panels A and B denoted when all participants received antibiotic treatment, if not treated earlier.

### Comparisons of MPO levels between LT-ETEC and (LT+ST)-ETEC strains

The pre-and post-challenge MPO levels were compared between LT+ST-ETEC (H10407)^25^ and LT-ETEC (LSN03-016011/A) strains. LT-ETEC induced comparable levels of MPO to (LT+ST)-ETEC challenge (LT-ETEC: overall 11-fold increase, MSDs: 12-fold increase, non-MSDs: 10-fold increase vs (LT+ST)-ETEC: overall 10-fold increase, MSDs: 20-fold increase, non-MSDs: 6-fold increase). MPO levels rose more rapidly following LSN03-016011/A challenge, peaking two days post-challenge (day 3), compared to (LT+ST)-ETEC, which peaked at three days post-challenge (Fig 2A). Non-MSDs induced by (LT+ST)-ETEC showed a gradual increase in MPO, while non-MSDs induced by LT-ETEC exhibited a rapid increase in MPO (Fig 2B). The challenge dose of (LT+ST)-ETEC ranged from 10^5^ to 10^8^ CFU, while the LT-ETEC dose was 5×10^9^ CFU.

**Fig 2.**
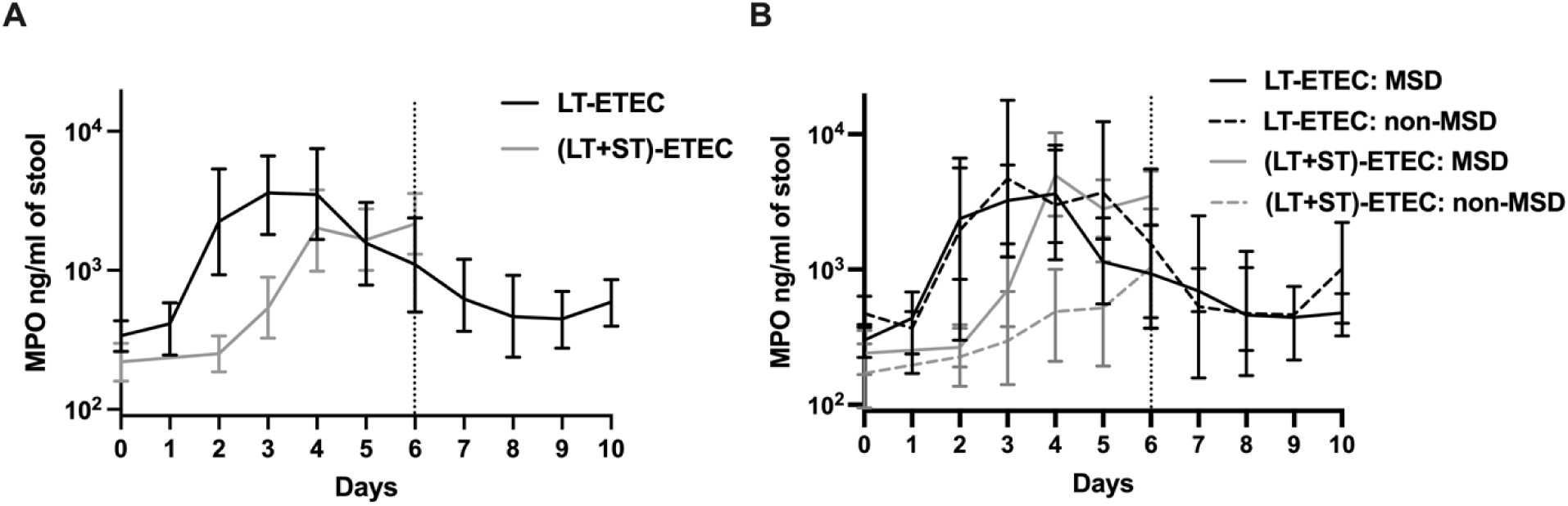
Comparisons of MPO levels between pre- and post-challenge with LT-ETEC (LSN03-016011/A) and (LT+ST)-ETEC (H10407) (log 10 scale).

Panel A, black line: Magnitude and kinetics of MPO levels among all participants infected by LT-ETEC; grey line: Magnitudes and kinetics of MPO levels among all participants infected by (LT+ST)-ETEC. Panel B, black solid line: Magnitudes and kinetics of MPO levels among participants with MSD infected by LT-ETEC; black dashed line: Magnitudes and kinetics of MPO levels among participants without MSD infected by LT-ETEC; grey solid line: Magnitudes and kinetics of MPO levels among participants with MSD infected by (LT+ST)-ETEC; grey dashed line: Magnitudes and kinetics of MPO levels among participants without MSD infected by (LT+ST)-ETEC. The dashed vertical line on Day 6 (120h post challenge) denotes when all participants were receiving antibiotic treatment, if not treated earlier.

### Kinetics of cytokine IL-1β after challenge

The geometric mean of fecal IL-1β increased significantly on days 2-7 after challenge, peaking on day 3 with a 107.86-fold increase over baseline (p=0.002), which decreased thereafter (Fig 3A). A similar pattern was seen among both participants with and without MSD. Those with MSD experienced 73.55-fold (p=0.009) increase on day 3 compared to baseline (Fig 3B).

**Fig 3.**
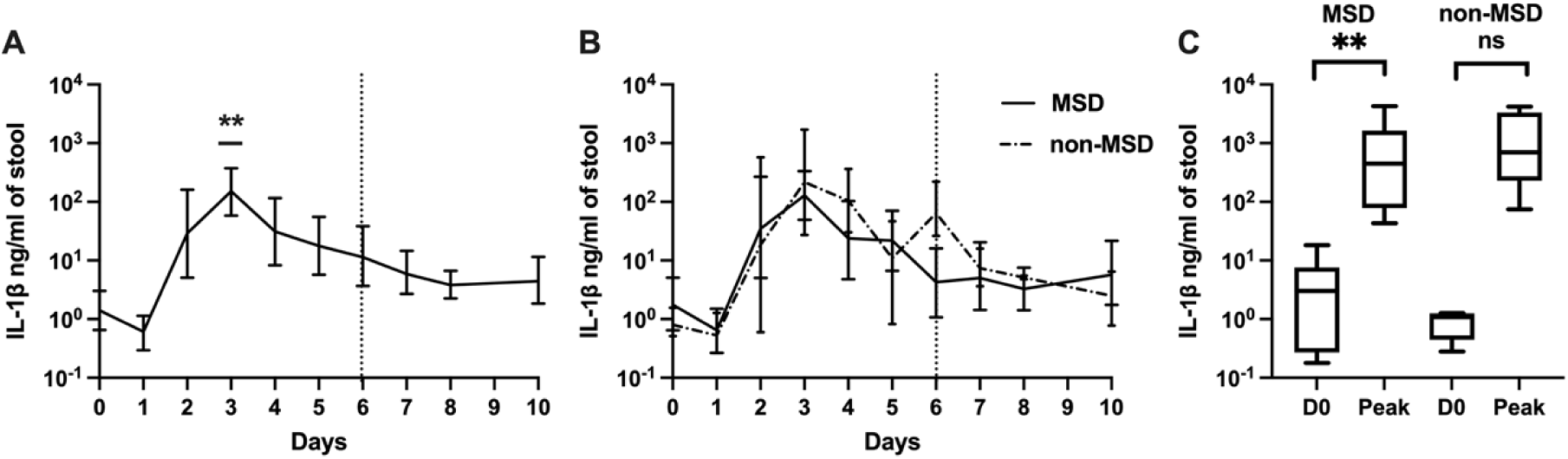
IL-1β levels pre- and post ETEC challenge. (log 10 scale).

The days when IL-1β levels peaked varied by participants, ranging from day 2 to day 6. The GM of the peak IL-1β on any day across all participants was 317.73-fold higher than baseline (p=0.001) and 221.55-fold higher among those with MSD (p=0.006) (Fig 3C). Baseline IL-1β levels were 2.21-fold higher in those with MSD compared to those without, though the difference was not statistically significant (p=0.479).

A. Magnitudes and kinetics of IL-1β levels among all participants. B. Magnitudes and kinetics of IL-1β levels based on clinical outcome following ETEC challenge. C. Comparison of baseline IL-1β and peak IL-1β levels on any day by clinical outcome. D0: day before challenge; Peak: peak IL-1β levels on any day. MSD: moderate to severe diarrhea; non-MSD: without moderate to severe diarrhea. p<0.01: **; not significant: NS. The dashed vertical line on Day 6 (120h post challenge) in panels A and B, denotes when all participants were receiving antibiotic treatment, if not treated earlier.

### Kinetics of cytokine IL-8 after challenge

Fecal IL-8 followed a similar kinetics as IL-1β among the participants after receiving the ETEC challenge (Fig 4). The GM of IL-8 peaked on day 3 with a 34.77-fold increase from baseline after the challenge (p=0.002). A significant increase was also found when comparing the GM of the peaks of IL-8 on any day following challenge to baseline among all participants (60.69-fold, p=0.001) and among those with MSD (57.79-fold, p=0.006). No significant association was observed between pre-challenge IL-8 levels and having MSD or non-MSD following challenge (MSD/non-MSD=0.93).

**Fig 4.**
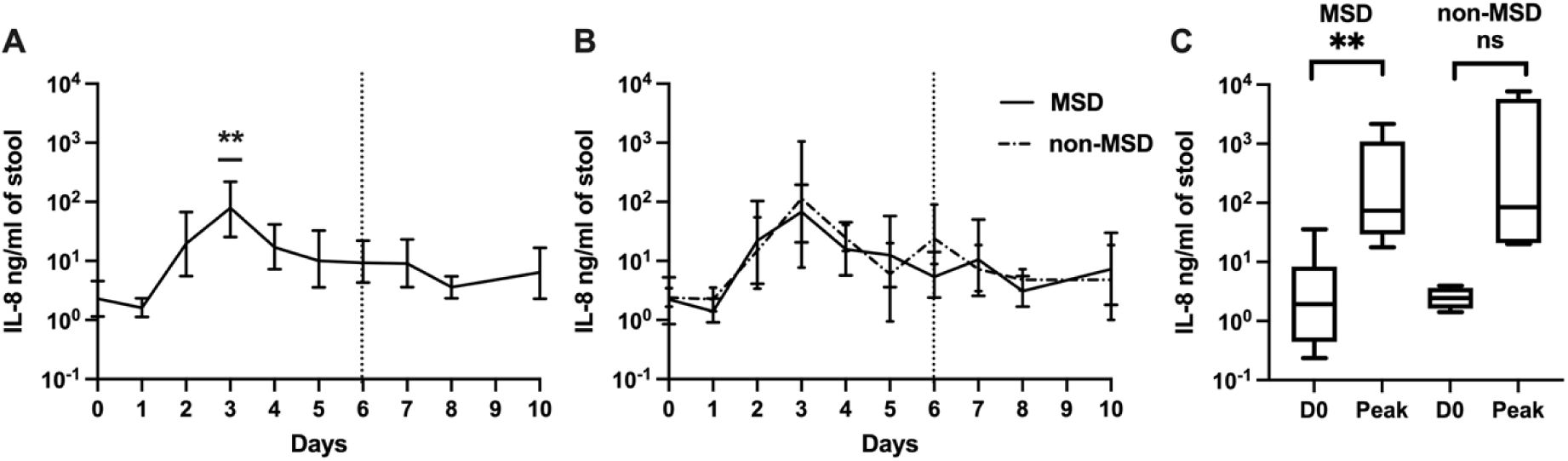
IL-8 levels pre- and post ETEC challenge. (log 10 scale).

A. Magnitudes and kinetics of IL-8 levels among all participants. B. Magnitudes and kinetics of IL-8 levels based on clinical outcome following ETEC challenge. C. Comparison of baseline IL-8 and peak IL-8 levels on any day by clinical outcome. D0: day before challenge; Peak: peak IL-8 levels on any day. MSD: moderate to severe diarrhea; non-MSD: without moderate to severe diarrhea. p<0.01: **; not significant: NS. The dashed vertical line at Day 6 (120h post challenge) in panels A and B denotes when all participants were receiving antibiotic treatment, if not treated earlier.

### Kinetics of other cytokines after challenge

The change in the kinetics of fecal cytokines IL-2, IL-4, IL-6, IL-10, IL-13, IL-17A, TNF-α, and IFN-γ following challenge are shown in S1 Fig. None of the eight cytokines demonstrated a >2-fold increase in geometric mean on any day following challenge compared to baseline. However, when comparing the GM of the peak values on any day to the baseline value, only IL-2 had a >4-fold difference (4.42-fold, p=0.003), (Peak GM=4.49 ng/ml, 95%CI: 2.88, 6.88).

Baseline titers of IL-2, IL-4, IL-6, IL-10, IL-17A, TNF-α, and IFN-γ were compared to determine if the pre-challenge cytokine levels were associated with the post-challenge diarrhea outcomes of ETEC challenge. Although not statistically significant, the GM of cytokines IL-10, IL-13, and IFN-γ on the day before the challenge among participants without MSD were 2.07-fold, 1.97-fold, and 1.67-fold higher, respectively than among those with MSD (Fig 5).

**Fig 5.**
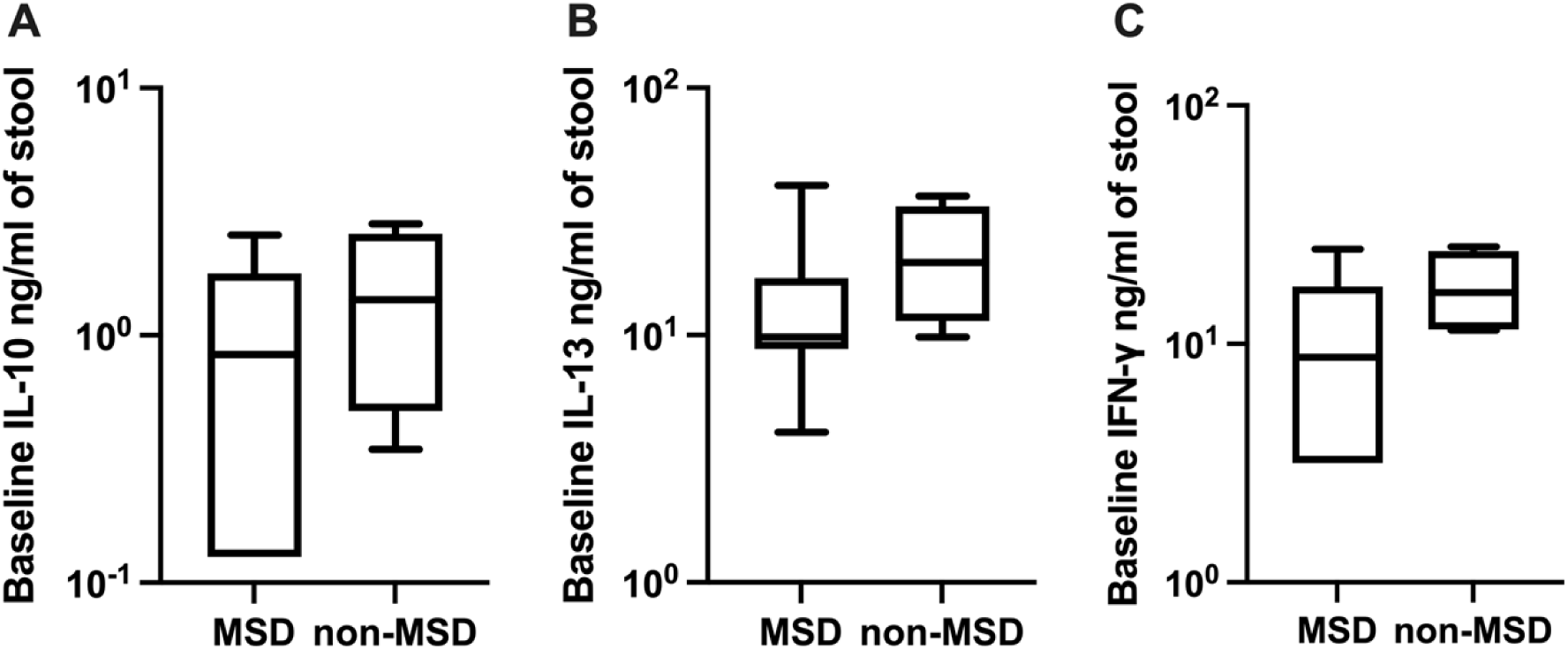
Pre-challenge levels of IL-10, IL-13, and IFN-γ among the participants with MSD or non-MSD. (log 10 scale). MSD: moderate to severe diarrhea; non-MSD: without moderate to severe diarrhea.

### Correlation between MPO and cytokines

A significant correlation was found among MPO, IL-1β, and IL-8. Peak level of MPO and peak fold change of MPO had a positive correlation with peak level of IL-1β (ρ =0.653, p=0.011; ρ =0.657, p=0.011). Similarly, the peak level and fold change of MPO were positively correlated with the peak level of IL-8 (ρ =0.675, p=0.008; ρ =0.653, p=0.011). The correlations between baseline, peak titer, and peak fold change of MPO and other cytokines among all participants were included in S1 Table.

### Correlation between inflammation and shedding

ETEC shedding in stool, which reflects colonization of ETEC in the intestine after the challenge, was monitored and evaluated until clearance of ETEC. The CFU of ETEC shedding ranged from 6.2X10^6^ to 4.8X10^9^. To minimize the impact of early antibiotic treatment, the CFU of shedding was compared after excluding one participant who received antibiotics 13.9 hours after challenge. No significant difference in shedding was observed 2 days post-challenge between those with MSD and those without. There was no significant correlation between the inflammation markers and ETEC shedding.

### Correlation between disease severity score and inflammation

Disease severity score post-challenge was inversely associated with baseline MPO (ρ =-0.577, p=0.024), baseline IL-4 (ρ =-0.550, p=0.042), baseline IL-6 (ρ =-0.588, p=0.027), baseline IL-10 (ρ =-0.656, p=0.011), baseline IL-13 (ρ =-0.650, p=0.012), peak IL-6 (ρ =-0.572, p=0.033), and peak IL-13 (ρ =-0.644, p=0.013).

### Association between MPO and immune responses to ETEC vaccine-specific antigens

A higher GM of peak MPO was observed among serum IgA responders and ALS responders to CS17 and CTB antigens (Fig 6).

**Fig 6.**
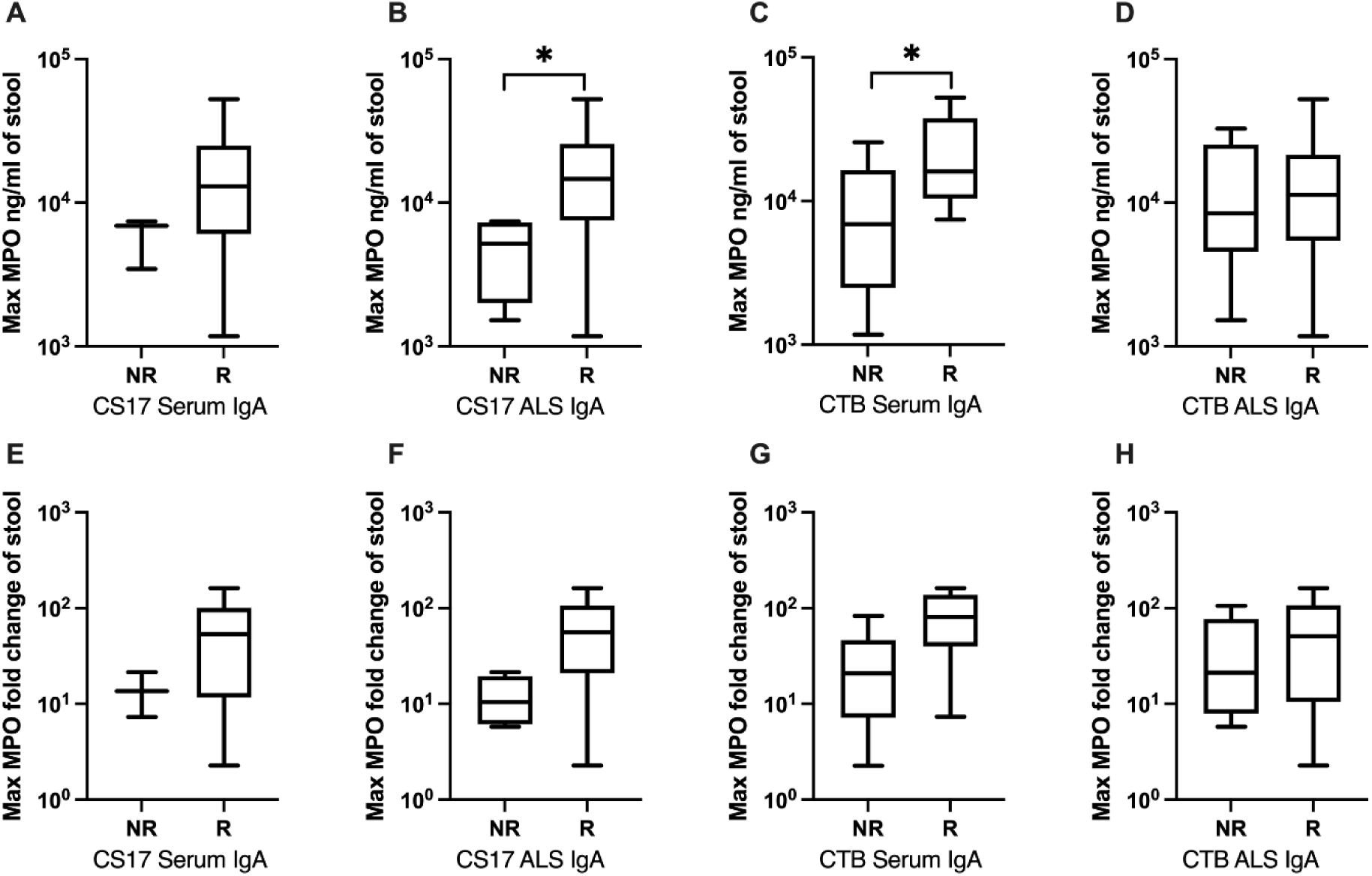
Post-challenge peak MPO levels and peak MPO fold change in responders and non-responders to CS17 and/or CTB following challenge. (log 10 scale). A, E: CS17 serum IgA; B, F: CS17 ALS IgA; C, G: CTB serum IgA; D, H: CTB ALS IgA; A, B, C, D: peak MPO level; E, F, G, H: peak MPO fold change; R: responders, NR: non-responders. p<0.05:*.

The peak MPO and peak MPO fold change among responders of CS17 serum IgA were 10688.1 (range: 1178-52500) ng/ml and 33.6 (range: 2.3-160.8) respectively, while the corresponding values among non-responders were 5615.9 (range: 3456-7424.2) ng/ml and 12.9 (range: 7.3-21.5) (1.9-fold difference, p=0.180; 2.60-fold difference, p=0.233).

For ALS IgA against CS17, the GM of peak MPO among responders was 12758.8 (range: 1178-52500) ng/ml, significantly higher than that among non-responders, which was 4053.1 (range: 1523.7-7424.2) ng/ml (3.15-fold difference, p=0.040). The GM of peak MPO fold change was 39.4 (range: 2.3-160.8) among the responders group, which was 3.75-fold higher than among non-responders [10.5 (range: 5.8-21.5), p=0.056].

Similarly, responders exhibited a higher peak and peak fold change of MPO compared to non-responders in CTB serum IgA. Responders had a peak MPO of 18267.9 (range: 7424.2-52500) ng/ml and fold change of 59.6 (range: 7.3-160.8), while non-responders had a peak MPO of 6033.2 (range: 1178-25687.5) ng/ml and fold change of 16.6 (range: 2.3-83.1) (p=0.0496, p=0.066). For CTB ALS IgA, the GM was higher in responders than in non-responders for peak MPO (9874 (range: 1178-52500) ng/ml vs. 8725.1 (range: 1523.7-32794.2) ng/ml, p=0.776) and peak MPO fold change (31.2 (range: 2.3-160.8) vs. 23.2 (range: 5.8-105.4), p=0.607). Additionally, peak MPO fold change was higher among responders (65.7, range: 25.6-160.8) of CS17 fecal IgA compared to non-responders (11.9, range: 2.3-67.4) (p=0.03). Baseline MPO was higher among non-responders (441.1, range: 264.2-1016.2) of CS17 fecal IgA compared to responders (225.8, range: 108.5-311.2) (p=0.017).

S2 Table includes the detailed results of the associations between MPO levels and seroconversions.

Seroconversion status was not significantly associated with clinical outcome (MSD vs non-MSD).

### Association between cytokines and immune responses to ETEC vaccine-specific antigens

The comparison of post-challenge cytokine levels by seroconversion status revealed several significant associations between responders and non-responders (Table 2). Since the concentration of cytokines varies, we tested different cut-off levels in this analysis, and the significant associations are included below. The detailed results are included in the S3 Table.

**Table 2.** Post-challenge peak cytokine levels by seroconversion status.

| Antibody | Cut<br>off | Cytokine | Non-responder | Responder | P<br>value |
| --- | --- | --- | --- | --- | --- |
| IL-1β | | | | | |
| CS17 serum IgA | 8 | IL-1β fold change | 121.8 (4.5-3331.1) | 1783.7 (266.3-15258.9) | 0.029 |
| CS17 ALS IgA | 16 | IL-1β | 80.0 (43.4-188.1) | 887.9 (74.8-4288.6) | 0.014 |
| CS17 ALS IgA | 16 | IL-1β fold change | 37.6 (4.5-357.8) | 745.9 (31.1-15258.9) | 0.036 |
| CS17 fecal IgA | 8 | IL-1β | 60.1 (43.4-84.4) | 757.3 (74.8-4288.6) | 0.024 |
| IL-2 |  |  |  |  |  |
| CTB ALS IgA | 4 | IL-2 | 8.1 (4.5-20.5) | 2.9 (1.3-11.1) | 0.013 |
| IL-4 |  |  |  |  |  |
| Total fecal IgA | 16 | IL-4 | 0.7 (0.2-1.6) | 1.8 (1.5-2.2) | 0.024 |
| CS17 serum IgG | 4 | IL-4 fold change | 4.1 (1.4-9.4) | 1.8 (1.0-3.3) | 0.045 |
| IL-6 |  |  |  |  |  |
| CS17 serum IgA | 8 | IL-6 | 1.8 (0.4-6.3) | 5.8 (3.5-9.2) | 0.007 |
| IL-8 |  |  |  |  |  |
| CS17 fecal IgA | 32 | IL-8 | 44.7 (17.7-145.6) | 1597.3 (350.5-7663.6) | 0.01 |
| CS17 fecal IgA | 32 | IL-8 fold change | 20.5 (2.8-205.5) | 1424.2 (574.1-5397.6) | 0.01 |
| CTB serum IgA | 2 | IL-8 | 52.7 (17.7-1124.7) | 788.8 (48.5-7663.6) | 0.019 |
| CTB serum IgA | 2 | IL-8 fold change | 15.8 (2.8-574.1) | 684.6 (102.1-5397.6) | 0.004 |
| CTB serum IgG | 2 | IL-8 fold change | 17.0 (2.8-574.1) | 332.3 (9.0-5397.6) | 0.02 |
| IL-10 |  |  |  |  |  |
| CS17 serum IgA | 8 | IL-10 fold change | 3.4 (1.0-14.3) | 1.3 (1.0-2.1) | 0.045 |
| CTB serum IgG | 2 | IL-10 | 1.3 (0.1-2.7) | 2.3 (0.8-3.4) | 0.043 |
| IL-13 |  |  |  |  |  |
| CS17 serum IgA | 4 | IL-13 | 8.9 (3.2-23.7) | 20.0 (11.4-35.3) | 0.029 |
| CTB fecal IgA | 8 | IL-13 fold change | 1.0 (1.0-1.0) | 1.7 (1.0-5.0) | 0.03 |
| IL-17A |  |  |  |  |  |
| CS17 serum IgA | 2 | IL-17A | 2.6 (1.6-7.4) | 8.7 (3.7-42.5) | 0.043 |
| CTB serum IgA | 2 | IL-17A fold change | 2.0 (1.0-4.9) | 4.8 (3.0-6.8) | 0.032 |
| TNF-α | | | | | |
| CS17 fecal IgA | 32 | TNF-α fold change | 1.8 (1.0-5.9) | 8.7 (3.6-18.3) | 0.019 |

Data were displayed as geometric mean (range). Only statistically significant comparisons ( $p < 0.05$ ) are shown.

### Cytokine levels positively associated with seroconversions to ETEC antigens

**IL-1β:** For CS17 serum IgA, the peak IL-1β fold changes were elevated among responders compared to non-responders. Similar trends were observed for CS17 ALS IgA where peak IL-1β levels and fold changes were significantly higher among responders. Moreover, peak IL-1β levels were higher among responders for CS17 fecal IgA.

**IL-6:** Peak IL-6 levels were significantly elevated among responders compared to non-responders for CS17 serum IgA.

**IL-8:** Peak IL-8 levels and fold changes were higher among responders than non-responders for CS17 fecal IgA and CTB serum IgA. Similar trends were observed for CTB serum IgG, where peak IL-8 fold changes were higher among responders compared to non-responders.

**IL-13:** Peak IL-13 levels were significantly higher among responders compared to non-responders for CS17 serum IgA. Moreover, for CTB fecal IgA, the fold change of peak IL-13 was higher among responders.

**IL-17A:** Responders showed notably higher peak IL-17A levels for CS17 serum IgA compared to non-responders. Similarly, for CTB serum IgA, responders had a significantly higher peak IL-17A fold change.

**TNF-α:** Peak TNF-α fold change was higher among responders than non-responders for CS17 fecal IgA.

### Cytokine levels negatively associated with seroconversions to ETEC antigens

**IL-2:** For IL-2, non-responders showed significantly higher peak levels than responders for CTB IgA in ALS.

### The association of cytokine levels and seroconversion status varied by antibody types

**IL-4:** Peak IL-4 level was significantly higher among responders for total fecal IgA than nonresponders. Conversely, IL-4 peak fold changes were elevated among non-responders compared to responders for CS17 serum IgG.

**IL-10**: While responders had a lower peak IL-10 fold change compared to non-responders for CS17 serum IgA, responders showed elevated peak IL-10 levels for CTB serum IgG.

Table 3 summarizes baseline cytokine levels associated with responders and non-responders across different antibody responses. Baseline IL-1β levels were significantly lower among responders for CS17 serum IgA compared to non-responders. A similar trend was observed for baseline IL-8 levels, which were lower among responders compared to non-responders for CTB fecal IgA.

**Table 3.** Pre-challenge cytokine levels by seroconversion status.

| Antibody | Cutoff | Cytokine | Non-responder | Responder | P value |
| --- | --- | --- | --- | --- | --- |
| CS17 serum IgA | 8 | IL-1β | 2.7 (0.2-18.2) | 0.4 (0.2-1.2) | 0.033 |
| Total fecal IgA | 8 | IL-2 | 0.8 (0.6-1.4) | 1.5 (1.3-1.8) | 0.045 |
| CTB fecal IgA | 32 | IL-8 | 2.8 (0.4-35.7) | 0.5 (0.2-1.3) | 0.033 |
| CS17 serum IgA | 8 | IL-10 | 0.4 (0.1-2.5) | 1.8 (0.9-2.8) | 0.023 |
| CS17 serum IgA | 4 | IL-13 | 6.0 (3.2-16.3) | 15.1 (7.1-25.6) | 0.032 |
| CS17 serum IgA | 2 | IL-17A | 1.4 (1.2-1.6) | 2.9 (1.2-7.7) | 0.049 |
| CS17 serum IgA | 8 | TNF-α | 1.5 (0.9-3.9) | 3.7 (1.8-7.2) | 0.029 |
| CS17 serum IgA | 8 | IFN-γ | 9.9 (4.1-35.4) | 22.7 (10.9-40.3) | 0.01 |

Data were displayed as geometric mean (range). Only statistically significant comparisons ( $p < 0.05$ ) are shown.

Conversely, baseline IL-10 levels, IL-13, IL-17A, TNF-α, and IFN-γ were significantly higher among responders than among non-responders for CS17 serum IgA. Baseline IL-2 levels were significantly higher among responders for total fecal IgA.

## Discussion

This study reports that infection with an ETEC strain producing only LT toxin induced significant intestinal inflammation in adult volunteers, both in those who had MSD and those who had mild to no diarrhea. Our study also showed intestinal inflammation might play a role in immune responses to ETEC-antigens. The CHIM study was done in a controlled inpatient environment, where all participants were dosed at the same time with a well-characterized LT-ETEC strain. This study provided a unique opportunity to measure intestinal inflammation solely induced by the infection with an LT-ETEC strain in a controlled environment in volunteers unlikely to be co-infected with other enteric pathogens.

MPO has been used in various studies as a biomarker for intestinal inflammation and was shown to be associated with growth failure and EED in children in LMICs^8,21,22^. ETEC was also shown to cause a significant increase in MPO in a mouse model^21^. In the MAL-ED study, infections with both LT-ETEC and ST-ETEC were associated with elevated MPO^8,22,44^. In this CHIM, we further underscored that LT-ETEC infection could induce intestinal inflammation. Inflammation in individuals with no or mild symptoms supports the potential for gut pathology secondary to asymptomatic infections with an LT-ETEC strain. Given that inflammation could be associated with growth failure, attention should be paid to ETEC-endemic areas where infected children may be overlooked because they are asymptomatic.

Notably, the level of MPO in both participants with and without MSD resulting from this ETEC strain producing only LT enterotoxin was comparable in many respects to the inflammation we observed in a previous study evaluating inflammation in participants experimentally challenged with H10407, ETEC strain producing both ST and LT enterotoxin^25^. Notably, the MPO response progressed more rapidly after LT-ETEC infection than after (LT+ST)-ETEC infection. However, the kinetics of the MPO response may reflect the differences in challenge dose and time to diarrhea onset for these two strains. At the LT-ETEC dose given, the time to first unformed stool in the diarrhea episode was approximately 22 hrs; whereas for (LT+ST)-ETEC, the time to first diarrheal stool was longer, ranging between 28-48 hrs^45^.

In our study, LT-ETEC induced significant levels of IL-1β and IL-8, which was also consistent with previous studies utilizing in vitro, murine, and porcine models. The kinetics of IL-1β and IL-8 had a positive correlation with the peak level and fold change of MPO. IL-1β and IL-8 are both pro-inflammatory cytokines produced primarily by activated macrophages, which explains their increase along with the inflammation biomarker MPO. However, other cytokines, including proinflammatory cytokines such as IL-6, IFN-γ, and TNF-α, or the anti-inflammatory cytokines IL-4, IL-10, and IL-13, although previously shown to change after ETEC infection in animal or cell culture models, didn’t significantly increase in this study^14,15,17,20,46,47^.

Notably, ETEC-induced intestinal inflammation was associated with immune responses to ETEC-specific antigens. IgA seroconversion in serum and ALS against CS17 and CTB was positively correlated with the peak levels of MPO. This may be due to participants with greater inflammation and symptoms tending to develop a stronger immune response to fight the infection. A similar positive relationship was found between MPO levels and serum titers against *Vibrio cholerae*^48^. Interestingly, in our previous study of ETEC H10407 expressing LT, ST, and CFA/I, inflammation was negatively associated with seroconversion to CFA/I IgA but, similar to this study, was positively associated with LT^25^. More studies with various combinations of ETEC toxins and CFs are needed to assess whether the association between inflammation and immune responses is antigen-specific.

This study also provides valuable insights into the cytokine responses associated with seroconversion status. Several pro-inflammatory cytokines, including IL-1β, IL-6, IL-8, IL-17A, and TNF-α, exhibited significantly higher peak levels or fold changes among the responders compared to non-responders. These cytokines have been found to show a synergetic increase; for example, TNF-α stimulates the production of IL-1β and IL-6, which may explain the consistent patterns observed among them^49^. ETEC LT has been demonstrated to induce a Th17 response, which thus produces IL-17A^50,51^. IL-17A is involved in the protection of the host against extracellular pathogens through the induction of sIgA^52–54^. In a previous study, LTB-specific IgA antibodies in ALS, but not plasma samples, were correlated with IL-17A in blood samples of adult Bangladeshi patients hospitalized with any ETEC diarrhea^55^. Collectively, these findings underscore the role of pro-inflammatory cytokines in driving immune responses and facilitating the clearance of ETEC. For anti-inflammatory cytokines, IL-10 showed distinct patterns depending on the antigen type, with the peak value lower in responders for CS17 antibody, but higher in responders for CTB antibody. This variation highlights the need for further research into antibody-specific IL-10 dynamics. IL-13 showed consistently elevated peak levels in responders for both antibody types. This suggests that IL-13 may play a dual role in modulating immune responses during seroconversion, possibly enhancing antibody production while also reducing excessive inflammation.

Besides the Th17 response, we found a combination of Th1 and Th2 responses. Among Th1 cytokines, IL-1β and TNF-α levels were higher in responders, while IL-2 levels were lower. For Th2 cytokines, IL-4 levels were lower in responders, IL-13 levels were higher, and IL-10 showed varied patterns. These observations align with a previous study on oral immunization of mice with LT-B, which demonstrated both Th1 and Th2 responses, with Th1 responses occurring earlier and Th2 responses emerging later. This temporal shift between Th1 and Th2 pathways may reflect a dynamic immune strategy during seroconversion, potentially to decrease immune-mediated pathology^56^.

We investigated if the levels of pre-challenge inflammatory markers were associated with protection against MSD and seroconversion status. Higher pre-challenge levels of pro-inflammatory IL-1β were seen among those with MSD, and higher pre-challenge levels of IL-1β and IL-8 were seen in non-responders, suggesting that high levels of these cytokines before challenge may dampen the acute response to antigenic stimulation and aggravate diarrhea severity (MSD)^57^. In contrast, higher pre-challenge levels of IFN-γ and anti-inflammatory IL-10 and IL-13 were seen among those protected from MSD and among responders. A higher pre-challenge level of IL-10 was also shown to be associated with protection against the (LT+ST)-ETEC challenge in our previous study^25^. Higher pre-challenge levels of IFN-γ have also been associated with a reduced risk of developing campylobacteriosis following an experimental challenge^58^. In addition to IL-10 and IL-13, higher baseline levels of MPO, IL-4, and IL-6 correlated with lower disease severity scores, indicating that these cytokines might play a role in protection against ETEC diarrhea. The baseline levels of cytokines IL-17A and TNF-α, although not associated with protection against MSD, were higher in responders, likely because of their distinct functions in promoting antibody responses. These findings suggest that the interplay between pro- and anti-inflammatory cytokines at baseline may play a critical role in determining immunity and clinical outcomes. Further studies are needed to study the mechanisms by which this nuanced balance of baseline cytokines influences immunity and protection from disease.

This study has several strengths. We used an experimental human challenge model to study intestinal inflammation induced by ETEC, which enabled us to rule out the interference of any co-pathogens. We showed that an ETEC strain that produces only LT toxin could cause significant inflammation, which is comparable to that caused by an ETEC strain producing both LT and ST toxins. We also showed that the intestinal inflammation caused by LT-ETEC was comparable between participants with and without MSD. We highlighted the potential role of inflammation in immune responses against ETEC. This study also has limitations. The MPO level may vary with stool consistency, especially in participants experiencing watery diarrhea, and result in a varied fecal level of proteins. In this study, we adjusted for the weight of stool but not for protein level. However, it was shown in a previous study that the additional protein standardization didn’t increase the accuracy of MPO beyond the stool weight standardization^59^. The sample size of this study was small. Although this was a controlled CHIM study with limited variations between participants compared to the studies done in LMICs, with the low number of participants without MSD and variability in host factors, some potentially significant differences between those with and without MSD could have been masked. Third, due to the unavailability of serum samples from early time points after the challenge, we were unable to test for systemic inflammation. Fourth, the observed relationship could be impacted by unmeasured confounders.

In conclusion, our findings underscored the negative impact of LT-ETEC in causing moderate to severe diarrhea and inducing significant intestinal inflammation in a healthy American adult population. Given the potential long-term impacts of ETEC-induced inflammation, such as growth faltering, LT-ETEC should be included while estimating ETEC disease burden and developing a plan for disease control. Our study reinforces the need for ETEC vaccines and other therapeutics to reduce the ETEC disease burden and improve survival and long-term developmental outcomes in children.

## Acknowledgments

We want to thank the volunteers who participated in this study.

## Funding statement

This study was funded by the National Institute of Allergy and Infectious Diseases (NIAID) of the National Institutes of Health (NIH) under Award Number AI1168316 (SC). The CHIM trial was funded by VALNEVA. The content of this article is solely the responsibility of the authors and does not necessarily represent the official views of the NIH, NIAID.

## Disclosure statement

The authors report there are no competing interests to declare.

## Data availability statement

The relevant data are included in the manuscript and supplement materials. Due to ethical considerations, the data supporting this research are not available for public sharing. Specific data might be provided upon request.

## Supporting information

**S1 Fig. Levels of cytokine IFN-γ, IL-13, IL-17A, TNF-α, IL-2, IL-6, IL-10, and IL-4 pre- and post ETEC challenge among all participants.**

**S1 Table. Correlation between MPO and cytokines**

**S2 Table. MPO levels by seroconversion status**

**S3 Table. Non-significant associations between cytokines and immune responses to ETEC vaccine-specific antigens**

**S4 Table. Associations between cytokines and immune responses to ETEC vaccine-specific antigens were analyzed using alternative seroconversion cutoffs**

## Notes

### Competing Interest Statement

The authors have declared no competing interest.

### Clinical Trial

NCT03576183

### Clinical Protocols

https://clinicaltrials.gov/study/NCT03576183#study-plan

### Funding Statement

Yes

### Author Declarations

The study was approved by the Bloomberg School of Public Health, Johns Hopkins University IRB (#00008616).

